# Ethnic Inequalities in Palliative Care Prescribing – A Rapid Systematic Review

**DOI:** 10.1101/2024.11.15.24317398

**Authors:** Tafadzwa Patience Kunonga, Eugenie Evelynne Johnson, Usvah Din, Elizabeth Westhead, Felicity Dewhurst, Barbara Hanratty

**Affiliations:** Population Health Sciences Institute, Newcastle University, Newcastle Biomedical Research Building, Newcastle upon Tyne, NE4 5PL, UK

**Keywords:** Palliative care, ethnic inequalities, prescribing, symptom management, high-income countries, rapid systematic review

## Abstract

**Context:** Effective palliative care involves managing the many symptoms commonly experienced towards the end-of-life. Appropriate prescribing is key to this care, and ethnic inequalities may lead to unequal treatment and poorer outcomes for minority groups. Understanding these disparities is critical to ensuring equitable care.

**Objectives:** To investigate ethnic inequalities in palliative care prescribing amongst adults residing in high-income countries.

**Methods:** The review was registered with the International Prospective Register of Systematic Reviews (PROSPERO) under registration number CRD42023476977. We conducted searches across three electronic databases (MEDLINE, Embase, and CINAHL) from January 2000 to November 2023. Screening, data extraction and quality assessment were conducted independently by two reviewers. Quality was assessed using various JBI Critical Appraisal tools. Due to the heterogeneity of included studies, a narrative review was undertaken without a meta-analysis.

**Results:** Out of 7880 references identified, 10 studies met the inclusion criteria, all conducted in the United States. Overall, five studies were deemed to be high quality and five were fair quality. The studies highlighted ethnic disparities in palliative care prescribing. Minority populations were less likely to receive pain management medications, particularly opioids, compared to non-Hispanic whites. Increased age, female gender, lower socioeconomic status, and place of residence were also related to differences in prescribing practices.

**Conclusions:** This rapid systematic review suggests that there are ethnic inequalities in palliative care symptom management prescribing, highlighting a possible gap in care for ethnic minority patients. Research beyond the USA is needed to understand if there are international disparities in palliative care prescribing.

**Key message:** This article reports a rapid systematic review on ethnic inequalities in palliative care prescribing for symptom control in high-income countries. Findings highlight a gap in care for ethnic minority patients. Further investigation is needed to understand how widespread disparities in palliative care prescribing are, across the world.

## Introduction

Prescribing in palliative and end-of-life care is increasingly complex. Longstanding problems include dynamic symptoms, individual variability, unlicensed medication use and polypharmacy (1). Advances in oncological and non-oncological disease management have resulted in patients living longer with increasingly complex symptoms and side effects. As the population ages, living with multimorbidity and organ dysfunction becomes increasingly common (2), both of which impact drug metabolism and efficacy. Tailoring prescriptions to individual needs requires careful assessment and personalised approaches. Furthermore, most people in the last year of life receive multiple medications for various conditions. Balancing symptom control while minimising adverse effects is challenging. Professionals must consider potential interactions and prioritize essential therapies; success relies upon effective utilisation of the expertise of a multi-disciplinary team (3).

Specialist pharmacists can play a critical role in optimizing medication management for individuals receiving palliative care (4). They may conduct thorough reviews of patients’ medication profiles, assessing appropriateness, interactions, and potential adverse effects (5). Regular reviews ensure the medication regimen aligns with the patient’s evolving needs. Collaborating with the multidisciplinary team, pharmacists have the expertise to tailor drug regimens for symptom relief. Effective symptom management enhances patients’ quality of life. In addition, pharmacists may educate patients, families, and other healthcare professionals about proper medication use. This includes deprescribing, dosing instructions, administration techniques, and managing side effects.

Ethnic differences in prescribing can arise due to inequalities in access, cultural beliefs, language barriers, and varying responses to medications (6). To tackle these issues, it is essential to address care inequalities and ensure healthcare providers undergo cultural competence training. Understanding patients’ cultural backgrounds helps tailor treatment plans and ensures respectful communication (7). Access to interpreters or bilingual healthcare professionals is vital. Clear communication about medications, side effects, and treatment options bridges language gaps. Involving patients and families in decision-making is essential. Healthcare professionals must understand their preferences, beliefs, and concerns (8). Respect for cultural practices related to health and healing is crucial. Additionally, relying on evidence-based guidelines and pharmaceutical expertise is essential. It is important to recognise that certain ethnic groups may have unique pharmacogenetic variations. By integrating these strategies, healthcare professionals can provide equitable and effective care, regardless of patients’ ethnic backgrounds.

Previous research highlights the need to address ethnic inequality in palliative care experiences, with systematic synthesis of evidence lacking to date (9–11). Addressing a critical gap in existing research, our rapid review aims to systematically synthesise evidence on ethnic disparities in palliative care prescribing amongst adults in high-income countries. This synthesis offers a foundation for informed decision-making and improved care. Our approach ensures a rigorous examination of the complexities and cumulative effects of social determinants, intersecting identities, and life-course influences on palliative care prescribing practices.

### Research aim

This rapid systematic review aims to explore ethnic inequalities in palliative care prescribing amongst adults residing in high-income countries.

#### Conceptual Framework Underpinning Research

This systematic review uses the Kunonga Framework (see Supplementary Table 1) developed specifically designed to address the complexities of health inequalities and inequities within the context of evidence synthesis (12). The Kunonga Framework builds upon the established PROGRESS-Plus framework by integrating a focus on intersectionality and the life-course perspective.

PROGRESS-Plus serves as an acronym for identifying and analysing the social determinants of health, standing for: Place of residence; Race/ethnicity/culture/language; Occupation; Gender/sex; Religion; Education; Socioeconomic status; and Social capital. The “Plus” element includes additional factors such as age, disability, and sexual orientation (13, 14).

The Kunonga Framework enhances the PROGRESS-Plus framework by reinforcing the critical distinction between health inequality and health inequity, emphasising that not all health differences are inherently inequitable. It advocates for the incorporation of intersectionality within evidence synthesis, which necessitates recognising how multiple social identities and factors such as race, ethnicity, gender, and socioeconomic status interact to create unique and compounded barriers to health (15, 16). Furthermore, the Kunonga Framework encourages integrating the lifecourse perspective in evidence synthesis, examining how life events and transitions influence outcomes (17, 18). This perspective will help researchers understand how socioeconomic status, cultural norms, and systemic factors shape ethnic inequalities in palliative care prescribing (18).

## Methods

This rapid systematic review was conducted in accordance with Preferred Reporting Items for Systematic reviews and Meta-Analyses (PRISMA) guidelines (19), with adaptations to expedite the process, including using a limited number of databases and restricting the search to English language studies, as described in rapid review guidelines (20). The review was registered with the International Prospective Register of Systematic Reviews (PROSPERO), (registration number: CRD42023476977).

### Search strategy

We developed search strategies for MEDLINE, Embase and CINAHL. The search was developed in MEDLINE and adapted for Embase and CINAHL. Medical Subject Headings (MeSH), keywords, and relevant terms, including synonyms for “palliative care,” “prescribing,” and “ethnic minorities,” were incorporated into the search strategy. Searches were conducted between January 2000 and November 2023 and were limited to studies published in English. Complete search strategies are provided in Appendices 1 to 3.

### Eligibility criteria

The full eligibility criteria for the review are presented in Table 1. Studies were included if they focused on adults receiving palliative care across various settings and examined ethnic inequalities in prescribing symptom relief medication. Palliative care is defined as a holistic approach to improving quality of life for patients with life-limiting illnesses by managing symptoms and addressing their physical, emotional, social, and spiritual needs (21). We defined inequality as measurable differences in palliative care prescribing among ethnic groups that do not consider fairness. Examples include variations in the types and dosages of medications prescribed or the frequency of pain management interventions. Inclusion criteria, predetermined in the protocol, specified studies from Organisation for Economic Cooperation and Development (OECD) high-income countries (22), to ensure comparability and maintain similarity in palliative care standards. Only English language studies published from 2000 onwards were considered to ensure that findings were relevant to current practices and reflect the latest research advancements and trends. Studies were excluded if they involved non-adult populations, were conducted in OECD medium or low-income countries, lacked a specific focus on symptom control medications, dated before 2000, or did not compare prescribing patterns across ethnicities. Abstracts, conference proceedings, dissertations, books, and other forms of grey literature were also excluded to ensure the inclusion of high-quality, peer-reviewed evidence.

**Table 1:** Eligibility criteria.

| **Criterion** | **Include** | **Exclude** |
| --- | --- | --- |
| Population | Adults aged 18+ with a life-limiting illness with a focus on ethnic minorities.<br>Receiving palliative care from generalists or specialists' providers.<br>Taking medicines for symptom control | Studies carried out on people below the age of 18.<br>People who are not receiving palliative care<br>People who are not taking medicines to control end-of-life symptoms |
| Intervention | Palliative care prescribing of symptom control drugs e.g., pain, agitation, nausea/vomiting | Non-prescribing practices or interventions.<br>Disease modifying drugs. |
| Comparator | Comparison of prescribing practices across different ethnic groups, e.g., access to pain medication or symptom management | No comparison across different ethnic groups. |
| Outcomes | Ethnic disparities in palliative care prescribing for symptom control | Outcomes not related to disparities in prescribing practices. |
| Study design | Randomised control trials, cohort studies, cross-sectional (retrospective or prospective). | Case studies, qualitative studies, reviews, editorials. |
| Setting | Studies conducted in Organisation for Economic Co-operation and Development (OECD) high-income countries. | Studies conducted in non-OECD countries or low- and middle-income countries. |
| Publication type | Peer-reviewed journal articles.<br>Articles published from 2000 onwards. | Abstracts only, conference proceedings, dissertations, books, grey literature.<br>Articles published before 2000. |
| Language | Studies published in English. | Studies published in languages other than English. |

### Study screening and selection

The identified studies underwent initial processing through EndNote 21 (23). Duplicates within the dataset were identified and removed. The remaining studies were imported into Rayyan, a web-based software for systematic reviews, to facilitate screening (24). To ensure systematic and consistent screening, criteria were developed based on the predetermined inclusion and exclusion guidelines. The criteria were piloted on 10% of the studies to refine their utility and efficacy prior to full-scale implementation. Titles and abstracts were assessed in Rayyan and categorised as ‘included’ or ‘excluded’ based on the criteria. Included studies were exported back to EndNote 21 for full-text examination. Both stages of selection were performed independently by two researchers, with any conflicts resolved through discussion.

### Data extraction

A data extraction form was created using Microsoft Excel to capture relevant study characteristics, participant demographics, and pertinent information. Extracted data included author details, publication year, country, participant age, study setting, study design, sample size, ethnic groups, underlying condition, treated symptoms, medications prescribed, inter-ethnic differences, study conclusions and, where reported, other PROGRESS-Plus factors. The extraction form underwent pilot testing with 20% of included studies to refine its structure. The remaining studies were extracted by single researchers, with another checking for accuracy. Any conflicts were resolved through discussion or by a third researcher.

### Critical appraisal

We used the JBI critical appraisal tools to systematically assess study quality and relevance (25). JBI tools were applied to various study designs, including Analytical Cross-Sectional Studies (26), Cohort Studies (26), and Case Control Studies (27). Critical appraisal was assessed independently by two researchers, with any conflicts resolved through discussion or by a third researcher. In this review, overall risk of bias was assessed based on the percentage of criteria fulfilled: 0-33% of criteria fulfilled indicated high risk of bias; 34-66% of criteria fulfilled indicated fair risk; and 67-100% of criteria fulfilled indicated low risk. No studies were excluded based on their overall risk.

### Data synthesis

Due to heterogeneity in the populations, outcomes and analysis methods across studies, we employed a narrative synthesis using the principles of Synthesis Without Meta-analysis (28). Each synthesis was grouped by PROGRESS-Plus domains and stratified by type of medication. In cases where the medication class was specified, we used the given classification. However, if the medication class was not explicitly stated, we described the medications based on their intended symptom management. A comprehensive list of medications used for each symptom is provided in Appendix 4. Where possible, adjusted analyses reported in the studies were prioritised in the synthesis over unadjusted analyses and frequency data (e.g. Chi^2^ tests) to account for the potential effects of confounding on effect estimates. We did not transform data to a standardised metric but used vote counting based on P values and 95% confidence intervals, where appropriate, to determine whether there was an association between ethnicity and prescribing practices. To assess whether there were associations between PROGRESS-Plus domains and palliative care prescribing, two reviewers (TPK and EEJ) independently interpreted each result and resolved conflicts through discussion. The narrative synthesis of medication use, and prescribing for ethnicity was the primary analysis, as it aligned most closely with the overall review question. We tabulated results and provided a brief narrative summary for each analysis. Due to variations in how ethnicity was reported across the original studies, this systematic review used the terminology and classifications from the primary sources. This ensured accuracy, transparency, and preserved the integrity of the original data without oversimplifying complex ethnic identities.

## Results

### Results of the search

The search identified 10,485 potentially relevant papers. Following deduplication in EndNote 21 and Rayyan, a total of 2605 duplicates were removed. Title and abstract screening was performed on 7880 records, with 7708 excluded. A total of 172 studies underwent full-text screening, and 162 were excluded. Ten studies met the inclusion criteria and were included in the review (29–38). The PRISMA flow chart (Figure. 1) summarises the study selection and screening process.

**Figure 1:**
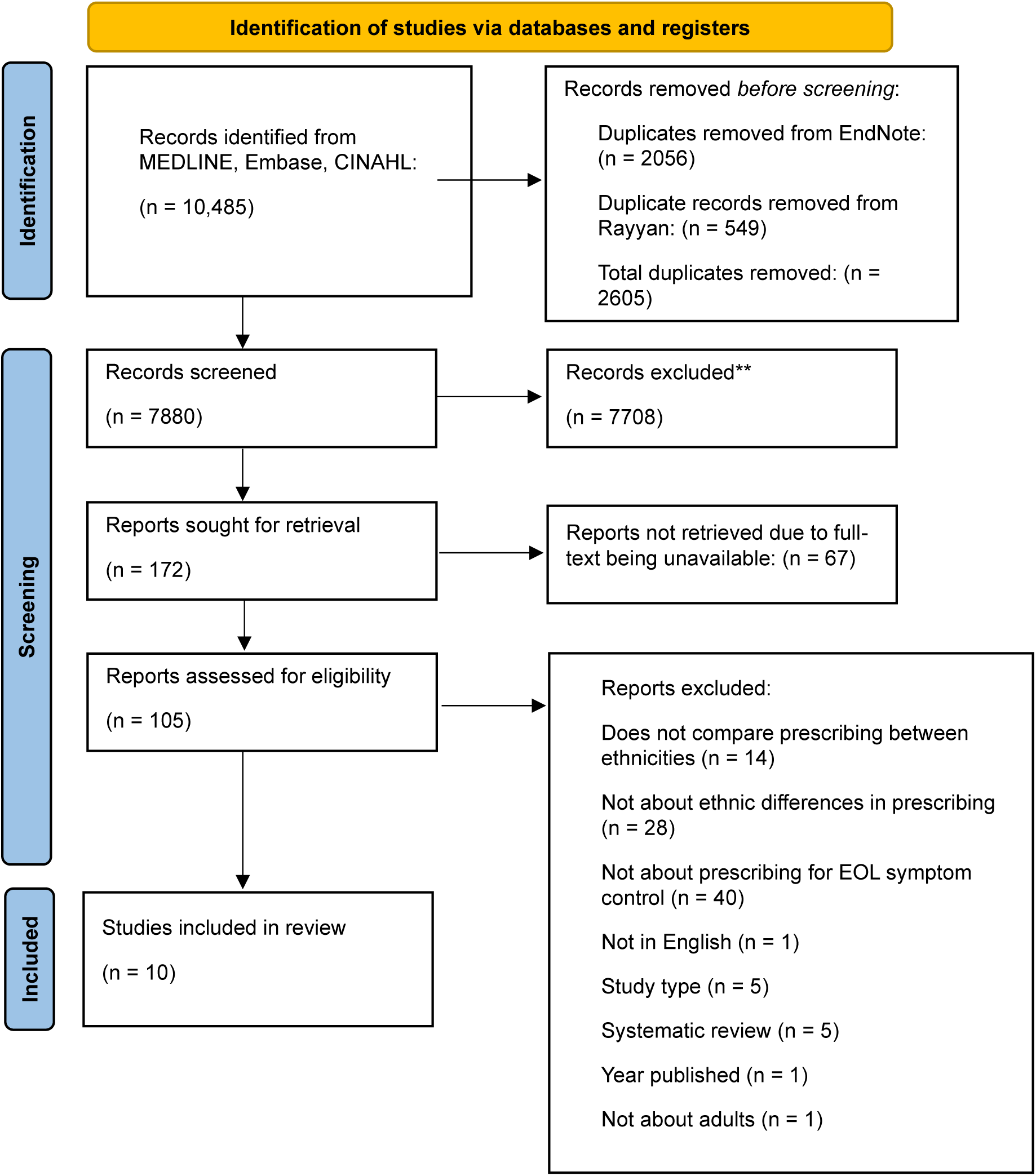
Flow diagram of study selection

### Study characteristics

Tables 2 presents an overview of individual study characteristics. Seven retrospective cohort studies (29, 31, 33, 35–38), two cross-sectional (32, 34), and one case control study,(30) all conducted in the United Staes of America (USA), were included. The data sources ranged from large-scale databases such as Surveillance, Epidemiology, and End Results (SEER)-Medicare and Medicaid claims files to specific healthcare settings like cancer centres and nursing homes. Sample sizes ranged from 64 (30) to 318,549 (30), participants, with a focus on older adults, typically aged over 65. Four studies focused on people with specific conditions: pancreatic cancer (29); lung cancer (37); ovarian cancer (35); and AIDS (36). The remaining six studies included people with a range of health conditions. Figure 2 gives an overview of all conditions within the included studies.

**Figure 2:**
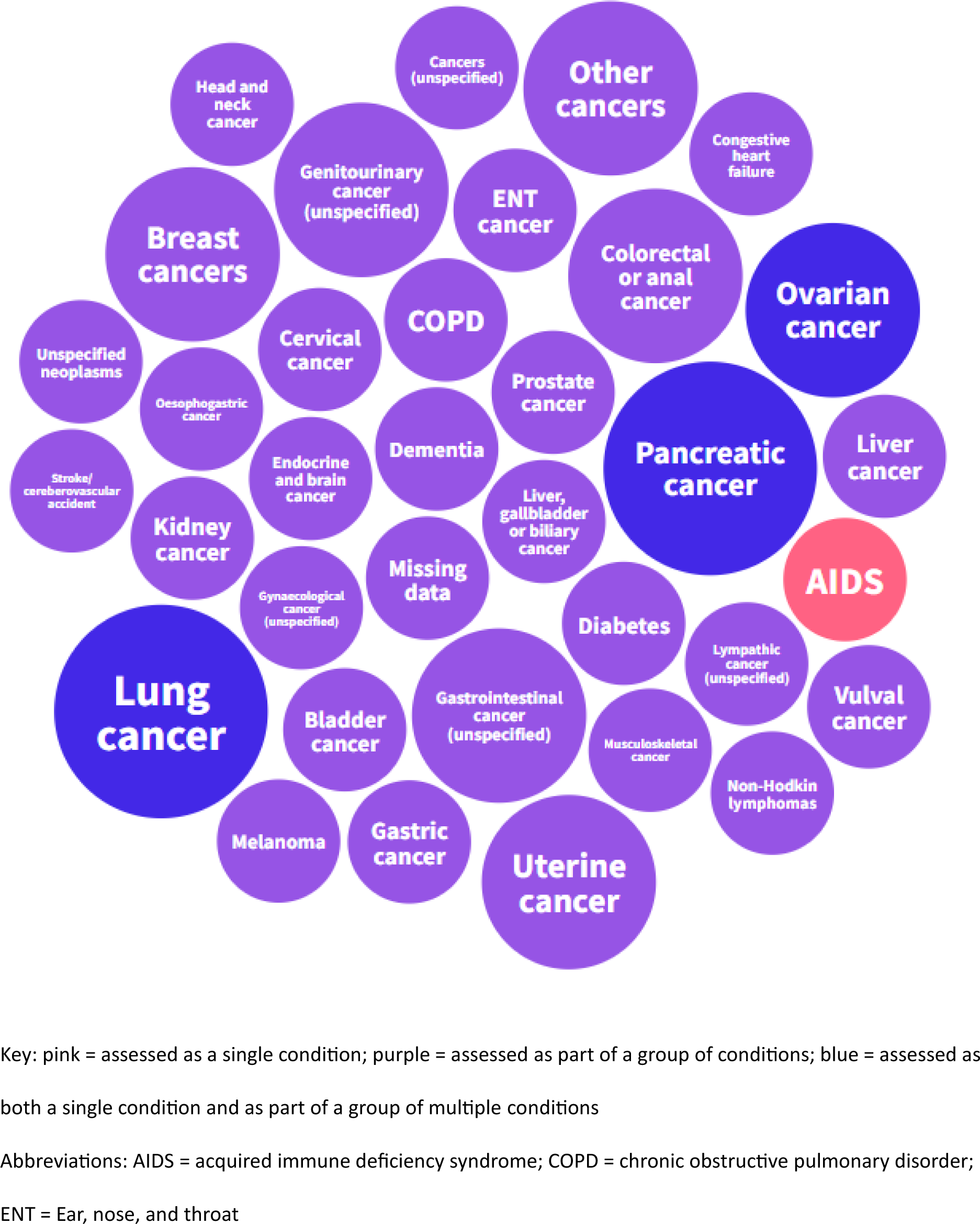
Overview of conditions from included studies

**Table 2:** Characteristics of included studies.

| Study ID | Country | Age | Setting | Study Design | Sample size | Ethnic Groups | Condition | Symptoms |
| --- | --- | --- | --- | --- | --- | --- | --- | --- |
| **Allen et al., 2023 (29)** | USA | 93% >65 years | SEER-Medicare-linked database from 2005 to 2017 | Retrospective cohort study | 74,309 | White (53,893); Black/African American (8,185); Hispanic (7,131); Asian (4,889); American Indian/Alaska Native (211) | Pancreatic cancer | Pain, psychiatric symptoms, depression, anxiety, muscle spasms |
| **Booker et al., 2020 (30)** | USA | >65 years | TRIP Cancer Pain Intervention from February 2007 to February 2010 | Case Control | 64 | African Americans (32); Caucasian Americans (32) | Cancer<br>Cancer diagnosis across all participants (n=64)<br>Gastrointestinal: 19; ENT: 17; Musculoskeletal: 6; Genitourinary: 5; Lymphatic: 4; Unspecified neoplasms: 2; Endocrine and brain: 1; Missing data: 10 | Pain |
| **Enzinger et al., 2023 (31)** | USA | >65 years | Centers for Medicare & Medicaid Services | Retrospective cohort study | 318,549 | White (272,358); Black (29,555); Hispanic (16,636) | Cancer<br>Cancer diagnoses across all participants (n=318,549) | Pain |
|  |  |  |  |  |  |  | Lung: 33.3%;<br>Colorectal or anal: 8%;<br>Pancreas: 6.8%;<br>Esophagogastric: 5%;<br>Liver, gallbladder, biliary: 5.2%;<br>Prostate: 6.6%;<br>Bladder: 2.6%;<br>Kidney: 2.4%;<br>Non-Hodkin lymphomas: 6%;<br>Acute leukaemias: 4.3%;<br>Breast: 5.8%;<br>Ovarian: 2.6%;<br>Uterine: 1.2%;<br>Melanoma: 1.4%;<br>other: 6% |  |
| **Haider et al., 2017 (32)** | USA | Median (range): 59 (51-67) | MD Anderson Cancer Center's outpatient palliative care clinic | Cross-sectional | 750 | White (529); Black (84); Hispanic (87); Other (50) | Cancer<br>Cancer types for total population (n=750)<br>Breast: 97;<br>GI: 193;<br>Genitourinary: 57;<br>Gynaecologic: 49;<br>Head and neck: 94;<br>Haematologic: 22;<br>Lung: 169;<br>Others: 69 | Pain |
| **Munir et al., 2023 (33)** | USA | 66+ | SEER-Medicare database from 2008 to 2016 | Retrospective cohort study | 48,631 | Black (5589), Asian (3089), Hispanic (5862), white (32,374) | Advanced gastrointestinal cancer<br>Liver: 6551;<br>Pancreas: 13,559;<br>Gastric: 5486;<br>Colorectal: 23,035 | Pain |
| **Reynolds et al., 2008 (34)** | USA | Median age (range): 81 (22-103) years | 12 nursing homes in central North Carolina from 2001 to 2004 | Cross-sectional | 1,133 | White 76.7%; ethnic minorities (not specified) | Dementia: 43.2%;<br>Stroke/Cerebrovascular accident: 28.4%;<br>Congestive heart failure: 23.7%;<br>Diabetes: 19.2%;<br>Chronic obstructive pulmonary disease (COPD): 9.1%;<br>Cancer: 4.8% | Pain |
| **Rolnick et al., 2007 (35)** | USA | Mean (range): 67 (20-79) years | Three large health maintenance organisations | Retrospective cohort study | 421 | White (348); African American (33); Asian (17); Hispanic (18); Native American (1); Unknown (4) | Epithelial ovarian cancer | Pain |
| **Sambamoorthi et al., 2000 (36)** | USA | 18+ | New Jersey AIDS Registry, Medicaid claims files for drug and non-drug services, and the Medicaid eligibility file | Retrospective cohort study | 2,131 | White (505); African American (1,228); Latino/Latina (389) | AIDS | Pain |
| **Saphire et al., 2020 (37)** | USA | Mean (SD): 76.8 (6.6) | SEER-Medicare database, January 1, 2008 to December 31, 2014 | Retrospective cohort study | 16,246 | White, non-Hispanic (13,077); Black, non-Hispanic (1,322); Hispanic (828); Asian, other (1,019) | Lung cancer<br>Non-small cell lung cancer: 12,898 (79.4%) | Pain, dyspnoea, emotional distress, fatigue, respiratory secretions, nausea, vomiting, anorexia |
| **Tabuyo-Martin et al., 2022 (38)** | USA | Mean (range): 58 (21 - 90) years | Sylvester Comprehensive Cancer Center | Retrospective cohort study | 186 | White (90); African American (46); Latino/Latina (42); Asian (6); Unknown (1) | Gynecological cancer<br>Cervix: 42 (22.6%);<br>Ovary: 70 (37.6%);<br>Uterus: 61 (32.8%);<br>Vulva: 13 (7.0%) | Pain, agitation, nausea/vomiting, lack of appetite, constipation |

**SEER: The Surveillance, Epidemiology, and End Results; TRIP: Translating Research into Practice; ENT: Ear, Nose and Throat; COPD: Chronic obstructive pulmonary disease; AIDS: Acquired immunodeficiency syndrome**

**Table 3:** Critical Appraisal of Retrospective Cohort Studies.

| Study ID | Question 1 | Question 2 | Question 3 | Question 4 | Question 5 | Question 6 | Question 7 | Question 8 | Question 9 | Question 10 | Question 11 | Overall Rating |
| --- | --- | --- | --- | --- | --- | --- | --- | --- | --- | --- | --- | --- |
| **Allen et al., 2023 (29)** | Yes | Yes | Yes | Yes | Yes | N/A | Yes | Yes | N/A | N/A | Yes | High |
| **Enzinger et al., 2023 (31)** | Yes | Yes | Yes | Yes | Yes | Yes | Yes | Yes | N/A | N/A | Yes | High |
| **Munir et al., 2023 (33)** | Yes | Yes | Unclear | Yes | Yes | Yes | Yes | Unclear | N/A | N/A | Yes | Fair |
| **Rolnick et al., 2007 (35)** | Yes | Yes | Yes | No | Unclear | Unclear | Yes | Yes | N/A | N/A | Unclear | Fair |
| **Sambamoorthi et al., 2000 (36)** | Yes | Yes | Yes | Yes | Yes | Yes | Yes | Yes | N/A | N/A | Yes | High |
| **Saphire et al., 2020 (37)** | Yes | Yes | Yes | Unclear | Yes | Yes | Yes | Yes | N/A | N/A | Yes | High |
| **Tabuyo-Martin et al., 2022 (38)** | Yes | Yes | Yes | No | No | Yes | Yes | Unclear | N/A | N/A | No | Fair |

**Table 4:** Critical Appraisal of Cross-Sectional Studies.

| Study ID | Question 1 | Question 2 | Question 3 | Question 4 | Question 5 | Question 6 | Question 7 | Question 8 | Overall Rating |
| --- | --- | --- | --- | --- | --- | --- | --- | --- | --- |
| Haider et al., 2017 (32) | Yes | Yes | Yes | Yes | Yes | No | Yes | No | High |
| Reynolds et al., 2008 (34) | No | Yes | Unclear | Yes | No | No | Yes | No | Fair |

**Table 5:** Critical Appraisal of Case Control Studies.

| Study ID | Question 1 | Question 2 | Question 3 | Question 4 | Question 5 | Question 6 | Question 7 | Question 8 | Question 9 | Question 10 | Overall Rating |
| --- | --- | --- | --- | --- | --- | --- | --- | --- | --- | --- | --- |
| Booker et al., 2020 (30) | No | Yes | Yes | Unclear | Unclear | No | No | Yes | Yes | No | Fair |

Symptoms targeted were predominantly pain, but also included psychiatric symptoms, dyspnoea, emotional distress, fatigue, respiratory secretions, nausea, vomiting, anorexia, constipation, and agitation.

In terms of PROGRESS-Plus variables, seven studies included race/ ethnicity/ culture/ language as a variable in their analyses, six included place of residence and gender/sex, and five included SES or personal characteristics. PROGRESS-Plus domains considered by the studies as part of their analyses are presented in Figure 3. Six studies considered intersectionality by adjusting for multiple PROGRESS-Plus variables in their analyses (29–31, 33, 36, 37). Only one study considered a lifecourse perspective in its analysis, accounting for the effects of persistent poverty on outcomes (33).

**Figure 3:**
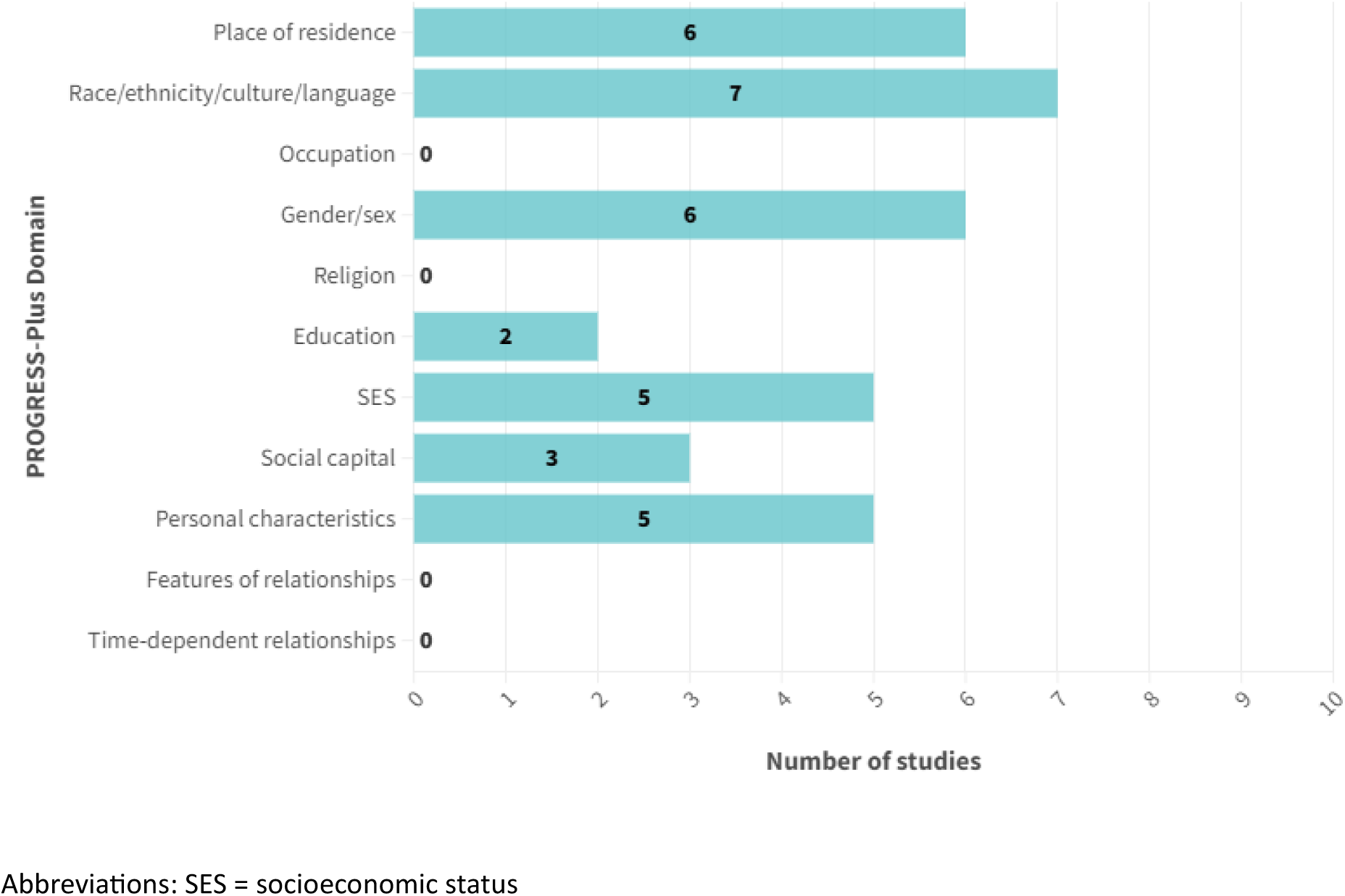
Number of studies including PROGRESS-Plus variables in analyses

### Critical appraisal of included studies

Overall, five studies were deemed to be high quality (29, 31, 32, 36, 37), with the remaining five studies judged as fair quality (30, 33–35, 38). For all studies judged as fair quality, there were some concerns surrounding whether the statistical analyses employed were appropriate and accounted for the effects of confounding. For the case-control study, there were concerns about the measurement of the exposure and comparability of cases and controls. For one cross-sectional study, there were concerns about the clarity of the eligibility criteria and method of exposure measurement.

### Synthesis of findings

#### Ethnicity

Nine studies reported on receipt of pain medications by ethic group (29, 31–38). One study reported on morphine equivalent daily dose (MEDD) across White, Hispanic, Black, and other ethnic groups, finding that Hispanic, Black, and Other racial groups were less likely to receive higher opioid doses compared to White patients (32). Although a statistically significant P value was provided, it was not clear between which groups this statistically significant association was. For the remaining eight studies, most studies suggested that people from ethnic minority groups may be less likely to receive any pain medication towards end of life, though one study suggested that Hispanic people may be more likely to receive pain medications (29), and two studies suggested no evidence of an association between ethnicity and prescribing (34, 38), (see Table 6 and Supplementary Table 2).

**Table 6:**
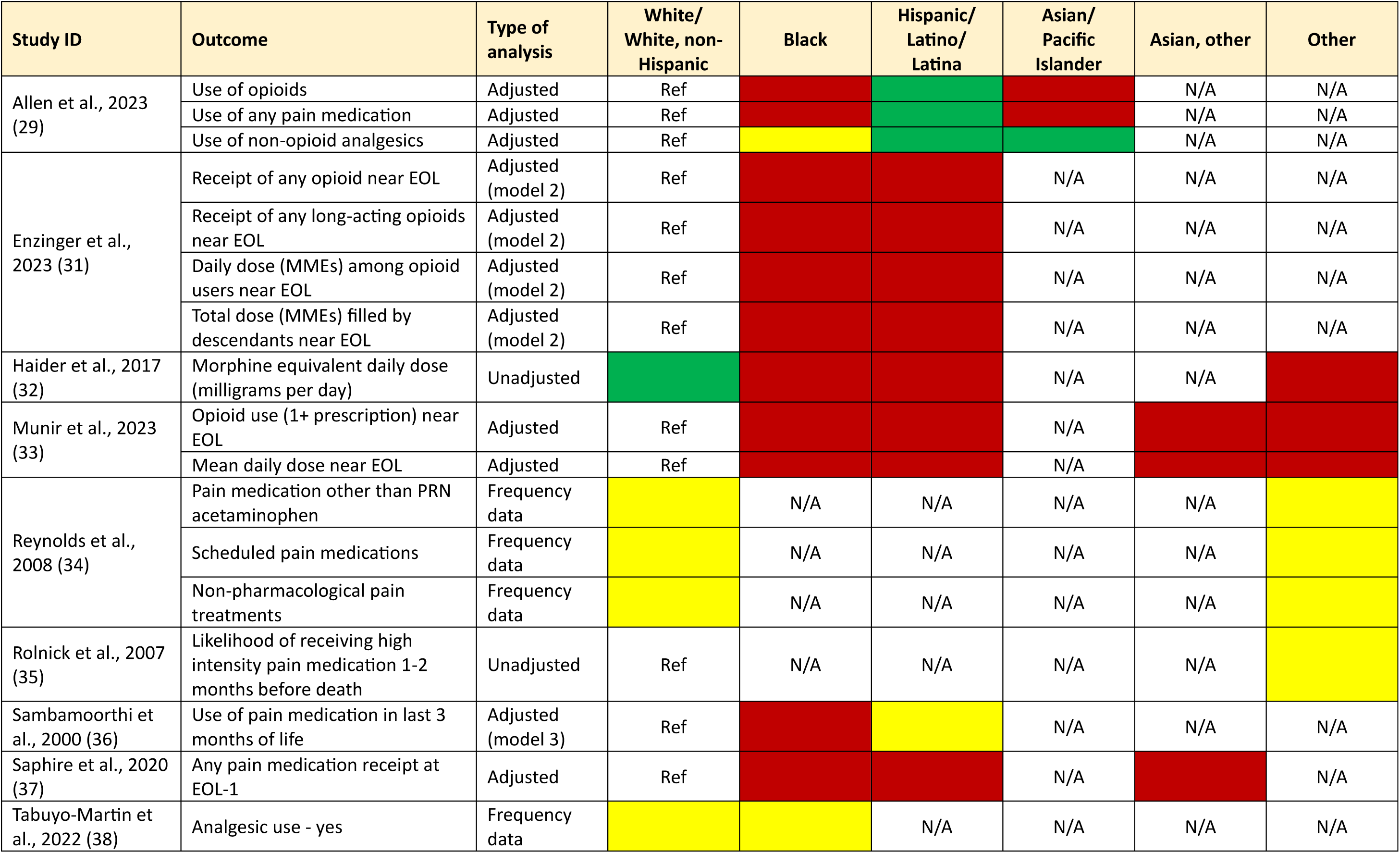
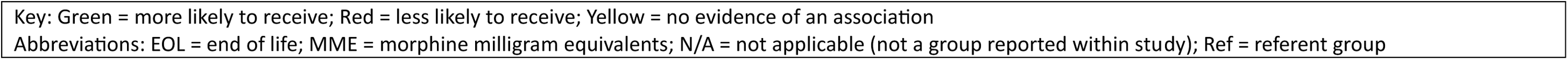
Ethnicity.

Two studies suggested that people from ethnic minority backgrounds were less likely to receive medications for emotional distress medications (37, 38), but the same two studies reported inconsistent associations for receipt of medications for nausea and vomiting. One study reported that people from ethnic minority backgrounds were more likely to receive medication for treating fatigue at end of life compared with white individuals (37). The same study suggested Black (non-Hispanic), Asian and other ethnic groups were less likely to receive medication for nausea at end of life compared with White individuals, though reported no evidence of an association antiemetics for Hispanic individuals compared with white individuals (37). The same study suggested that Black non-Hispanic and Asian individuals may be more likely to receive medications for anorexia at end-of-life compared to White individuals, while reporting no evidence of a discrepancy between prescribing for White and Hispanic individuals (37). One study reported that people from ethnic minorities may be less likely to be prescribed antidepressants, antipsychotics and anxiolytics than White individuals (29). Finally, one study suggested that Black individuals may be more likely to use appetite stimulants compared to White individuals (38).

#### Influence of other PROGRESS-Plus factors

Across the studies, various PROGRESS-Plus factors, including gender/sex, place of residence, age, socioeconomic status, and social status, were examined for their influence on access to palliative care and symptom management both in combination with and independent from ethnicity.

#### Gender

Five studies analysed receipt and use of medication towards end of life by gender while adjusting for the potential confounding effects of ethnicity (see Supplementary Table 3) (31–33, 36, 37). With regards to receiving pain medications, there were inconsistent results across the five studies. Two studies suggested that men were more likely to be prescribed or use pain medication (31, 32), while three suggested women were more likely to use pain medication (33, 36, 37).

Only one study assessed receipt of medications for anorexia, dyspnoea, emotional distress, fatigue and nausea one month before end of life (37). The study suggested that females were less likely to receive medications for anorexia and fatigue compared with males, but may be more likely to receive medications for dyspnoea, emotional distress and nausea (37).

#### Place of residence

Four studies analysed receipt and use of medication towards end of life by place of residence while adjusting for the potential confounding effects of ethnicity (see Supplementary Table 4) (31, 33, 36, 37). Results from these four studies assessing the likelihood of receiving pain medications were inconsistent. One study suggested that there may be no evidence of an association between place of residence and prescribing of medications for dyspnoea and emotional distress, but indicated that people living in less urban areas may be more likely to receive medications for anorexia and nausea (37). The same study suggested that urban residents were less likely to receive medications for fatigue compared with those living in a large metropolitan statistical area (37).

#### Personal characteristics: age

Two studies analysed receipt and use of medication towards end of life by age while adjusting for the potential confounding effects of ethnicity (see Supplementary Table 5) (33, 37). Both studies suggested that receipt of pain medication became less likely with increasing age (33, 37). One study suggested that there may be no evidence of an association between age and receipt of medications for anorexia, while those older than 66-69 may be more likely to receive medications for emotional distress and nausea. However, the same study presented inconsistent results across age groups in terms of receipt of medications for fatigue and dyspnoea (37).

#### Personal characteristics: age at diagnosis

One study assessed receipt and use of medication towards end of life by age at diagnosis while adjusting for the potential confounding effects of ethnicity (36), (see Supplementary Table 6), suggesting no evidence of an association between age at diagnosis and the use of pain medication in the last 3 months of life across all age groups (36).

#### Socioeconomic status

Four studies analysed receipt and use of medication towards end of life by SES while adjusting for the potential confounding effects of ethnicity (31, 33, 36, 37). SES was assessed using heterogenous measures across the four studies, while two of the studies assessed SES using two different measures (31, 33). Results of these studies by SES are presented in Supplementary Table 7.

#### Social capital

One study assessed receipt and use of medication towards end of life by a measure of social capital (marital status) while adjusting for the potential confounding effects of ethnicity (37), presenting inconsistent results (see Supplementary Table 8). The study suggested that unmarried individuals may be less likely to receive medications for anorexia, emotional distress, and nausea, compared to married individuals but there was no evidence of an association between marital status and receipt of medications for dyspnoea, fatigue, or pain (37).

#### Lifecourse Perspective

Only one study incorporated the lifecourse perspective in analysing ethnic inequality in palliative care prescribing (33). The study found that Black [adjusted odds ratio (aOR) = 0.84; 95% confidence interval (CI): 0.79–0.90], Asian (aOR = 0.86; 95% CI: 0.79–0.94), and Hispanic (aOR = 0.90; 95% CI: 0.84–0.95) patients with advanced gastrointestinal cancer had lower odds of filling an opioid prescription compared to White patients (33). Additionally, these groups received lower daily doses, with Black patients receiving 16.5 percentage points less and Hispanic patients 19.1 percentage points less. Disparities were somewhat reduced for Asian and Hispanic patients in high-poverty areas but worsened for Black patients (33).

## Discussion

### Summary of Main Findings

This rapid systematic review identifies ethnic disparities in palliative care prescribing among adults in high-income countries, with these disparities being further exacerbated by intersecting factors such as gender, place of residence, age, socioeconomic status, and social capital. However, it is important to acknowledge that the robustness of these findings is limited by the relatively small number of studies included in this review. a. In addition, the generalisability of the results is constrained by the fact that all the studies were conducted in the US, which may limit the applicability of the findings to other contexts.

Our review indicates that certain ethnic groups, including Black, Asian, and Hispanic patients, may be less likely to receive pain, emotional distress, and nausea medications, with particularly lower odds observed in opioid prescriptions (33, 37, 38). Gender further compounds these disparities, as women from these ethnic groups often receive less pain management (36, 37). Additionally, place of residence plays a crucial role, with minorities in less urbanised areas facing heightened barriers, while urban residents encounter distinct challenges (37). Age also intensifies these disparities, particularly among older ethnic minority patients, who are less likely to receive appropriate palliative care (33, 37). Socioeconomic status was found to interact with ethnicity to either mitigate or exacerbate these disparities, with some alleviation seen in high-poverty areas for Asian and Hispanic patients, although conditions worsen for Black patients (37). Furthermore, social capital, such as marital status, appears to influence care receipt, with unmarried individuals generally receiving less comprehensive treatment (37). The incorporation of a lifecourse perspective, though represented by a single study in this review, revealed how these disparities not only persist, but can worsen over time (33).

### Comparison with other work

This rapid systematic review adds to growing reports exploring inequalities in in prescribing across the life course, as well as in the provision of palliative care. Previous research has reinforced the need to reduce inequalities in prescribing, especially for people from ethnic minority backgrounds. One systematic review found significant racial disparities in pain treatment (39). Specifically, Black/African Americans had a lower likelihood of receiving opioid prescriptions for non-traumatic/non-surgical pain and Hispanics/Latinos also faced disparities for non-traumatic/nonsurgical pain, although not for traumatic/surgical pain (39). However, another review reported mixed results on racial and ethnic disparities in pain management, with over half of the studies showing no significant differences in pain medication receipt (40). These mixed findings are consistent with our review, which also observed variability in the extent of ethnic disparities, particularly in pain management and access to medications, however further research is needed to explore these patterns globally.

### Strengths and limitations

This rapid systematic review adhered to PRISMA guidelines, ensuring a robust and transparent methodological approach (19). The use of an intersectional lens and lifecourse perspective, as outlined in the Kunonga et al. framework, further strengthens the analysis by providing a nuanced understanding of how social determinants influence disparities in palliative care prescribing. However, this review is not without limitations. The search strategy was limited to three databases, which, although commonly used in health research, may not have captured all relevant studies. Expanding the search to additional databases or grey literature could have provided a more comprehensive evidence base. Although screening, data extraction, and critical appraisal were done independently by two reviewers, a single reviewer screened most studies in the initial stage. This may have increased the risk of bias or missed studies, despite efforts to minimize this through the double-checking of 20% of the studies by a second reviewer. Regarding the synthesis, we did not transform data to a standardised metric to enable possible comparison between studies. As a result, we relied on narrative synthesis, which, while appropriate for the heterogeneity of the included studies, limits the ability to quantify the overall effect of ethnic inequalities in palliative care prescribing. This may also affect the generalisability of the findings, as differences in study methodologies and outcomes could obscure potential patterns or associations.

### Implications for practice

Pharmacists play a pivotal role in optimizing medication management in both generalist and specialist providers of palliative care (41). Efforts to integrate pharmacists into multidisciplinary care teams in hospices, care homes and other settings, may help mitigate disparities in access to expert pharmacy services and prescribing, particularly for ethnic minority groups (42). Pharmacists’ expertise in medication optimization, can enhance palliative care for all patients (43). Furthermore, pharmacogenetic research highlights ethnic variations in drug responses, necessitating individualised prescribing approaches under expert supervision (44). However, underrepresentation of ethnic minorities in clinical trials limits our understanding of optimal medication use in palliative care (44). Future research focusing on personalized medicine could improve symptom control and align care with patient preferences (45).

More generally, there is increasing acknowledgement of the need to improve equity in palliative care provision, particularly for people from ethnic minority communities. This review suggests that one way to tackle inequities is to target discrepancies in prescribing practices for people with life-limiting illnesses. However, a multi-faceted approach is required to address this broad ranging and complex issue. Cultural competence amongst health and social care professionals is key, so that understanding and respect for the different influences on care preferences and needs becomes the norm.

### Implications for research

Incorporating an intersectionality lens (16, 46), and a lifecourse perspective (18), in future research is essential for advancing our understanding of ethnic disparities in palliative care prescribing. By examining how ethnicity intersects with other social determinants such as gender, age, socioeconomic status, and place of residence, researchers can better identify the compounded effects these factors have on prescribing in palliative care. The lifecourse perspective offers valuable insights into how these disparities develop and persist over time, highlighting the cumulative impact of social inequities on access to and quality of palliative care. However, the concentration of research in the US raises critical questions about what is happening in other countries. Additional research is required to explore potential disparities in palliative care prescribing within a broader, global context. Using methodologies, such as multilevel modelling (47), that can effectively capture these complex interactions will be critical for developing targeted interventions aimed at reducing inequities and improving care for underrepresented and vulnerable populations (18).

## Conclusion

While medicines are crucial for relieving symptoms in palliative care, our findings suggest potential disparities in prescribing practices, raising concerns about equitable access to palliative care across ethnicities. However, the concentration of research in the US raises critical questions about what is happening in other countries. Further research is needed to investigate potential disparities in palliative care prescribing in a broader, international context.

## Supporting information

PRISMA Checklist

## Disclosure/Conflict of Interest

The authors declare that they have no known competing financial interests or personal relationships that could have appeared to influence the work reported in this paper.

## Data Availability statement

All data produced in the present work are contained in the manuscript.

## Acknowledgements

None

## Funding

This research is funded through the National Institute for Health and Care Research (NIHR) Policy Research Unit in Older People and Frailty (funding reference PR-PRU-1217-2150). As of 01.01.24, the unit has been renamed to the NIHR Policy Research Unit in Healthy Ageing (funding reference NIHR206119). The views expressed are those of the author(s) and not necessarily those of the NIHR or the Department of Health and Social Care.

## Appendices

## Appendix 1 MEDLINE search strategy

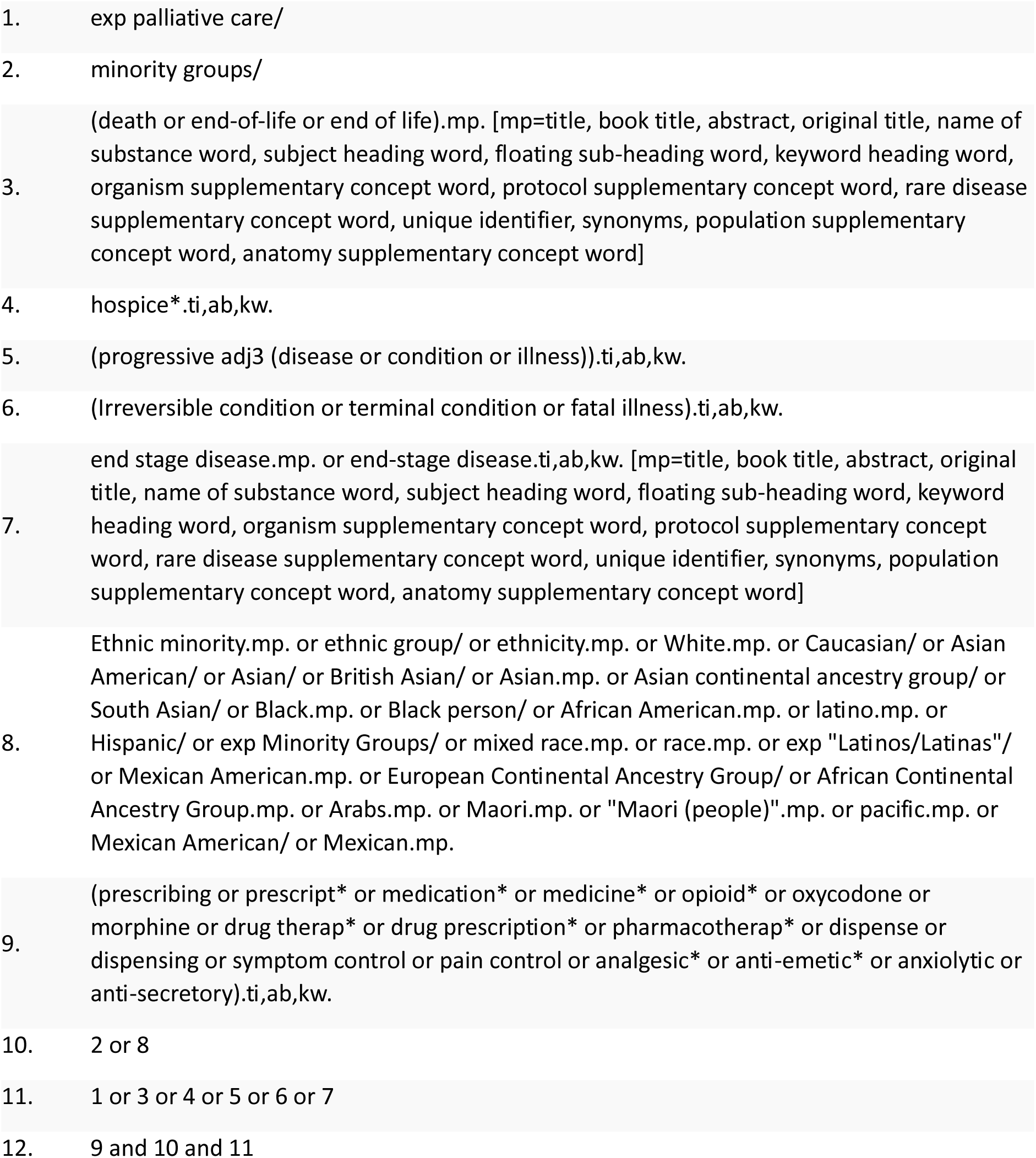

## Appendix 2 Embase search strategy

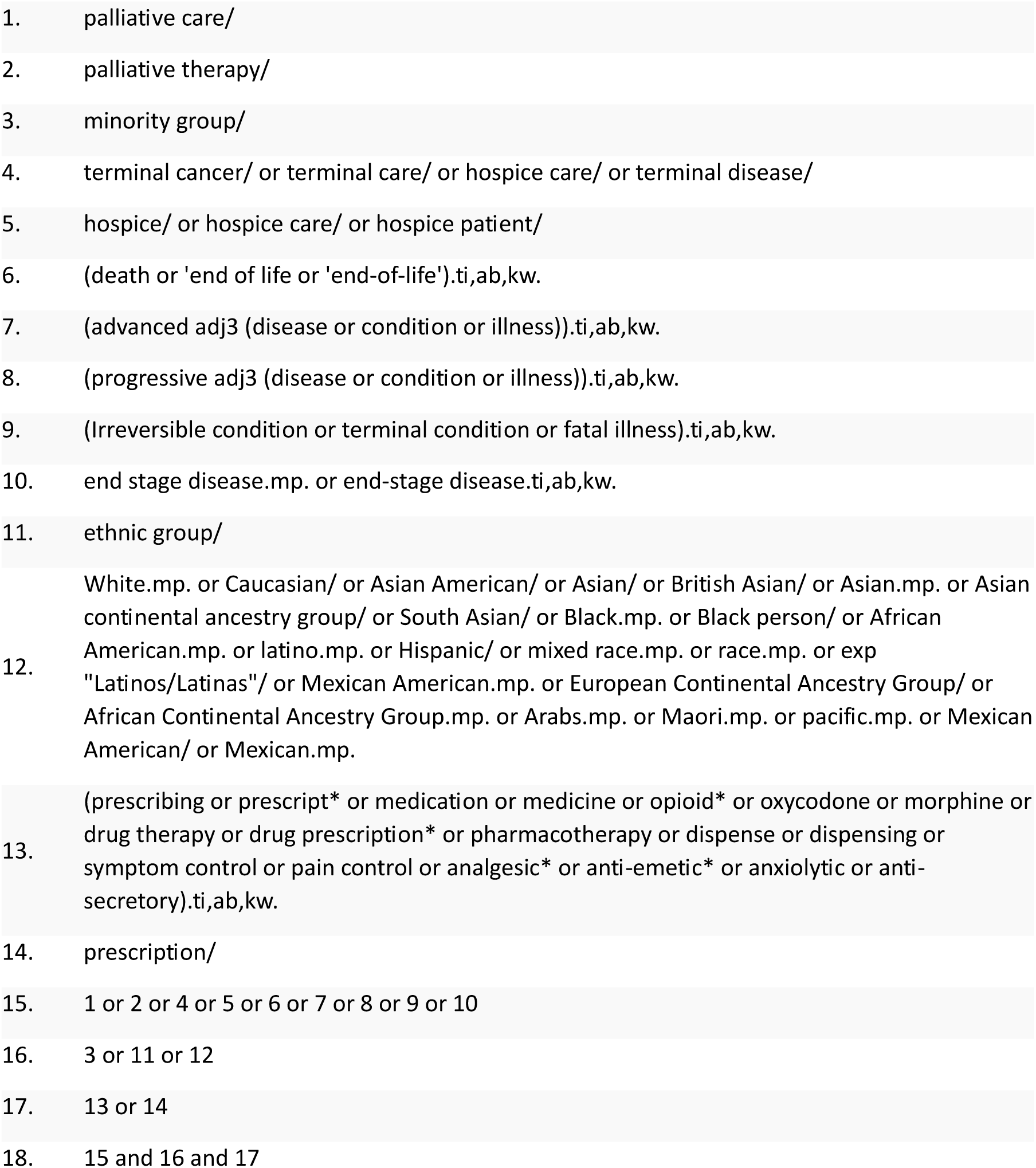

## Appendix 3 CINAHL search strategy

Appendix 3: CINAHL search strategy
|  |  |
| --- | --- |
| 1. | (MH "Palliative Care") OR "terminal care" or "end of life" or "end-of-life" or death or dying or hospice* OR (MH "Life Support Care") OR "advanced disease" or "advanced condition" or "advanced illness" OR "progressive disease" or "progressive condition" or "progressive illness" OR "irreversible condition" or "terminal condition" or "fatal illness" OR "end stage disease" or "end-stage disease") |
| 2. | ((MH "Minority Groups") OR "ethnic groups" or race or ethnicity OR white or Caucasian or "Asian American" or Asian or Black or "British Asian" or "South Asian" or "African American" or Latino or Latina or Mexican or African or Arabs or Maori or pacific) |
| 3. | ((MH "Pain Management") OR prescribing or prescription* or prescrib* or medication* OR opioid* or "pain management" or oxycodone or morphine or pharmacotherap* or dispens* or "symptom control" or "pain control" or analgesic* or anti-emetic* or anxiolytic or anti-secretory) |
| 4. | 1 AND 2 AND 3 |

## Appendix 4 List of medications used for each symptom in the study by Saphire and colleagues (37)

Appendix 4 :List of medications used for each symptom in the study by Sapphire and colleagues (37).
| Condition | Medications |
| --- | --- |
| **Pain** | Opioids, Nonopioid Analgesics, Adjuvants |
| **Emotional Distress** | Antidepressants, Anxiolytics/Sedatives, Antipsychotics |
| **Fatigue** | Transfusions, Erythropoietin-Stimulating Agents (ESAs), Methylphenidate |
| **Dyspnoea** | Inhaled Bronchodilators, Inhaled Corticosteroids, Diuretics, Oral COPD/Asthma Medications, Secretion Management Medications |
| **Anorexia** | Appetite Stimulants |
| **Nausea/Vomiting** | Serotonin (5-Hydroxytryptamine 3) Antagonists, Neurokinin-1 Antagonists, Prochlorperazine/Promethazine, Cannabinoids, Metoclopramide |

## Notes

### Competing Interest Statement

The authors have declared no competing interest.

