## Supplementary Files for "Ethnic Inequalities in Palliative Care Prescribing – A Rapid Systematic Review"

### Supplementary Table 1: The Kunonga Framework for Extending Existing Approaches in Inequality/Inequity-Focused Evidence Syntheses

| Stage | Component | Description |
|---|---|---|
| Protocol Stage | Define Key Terms and Scope | In addition to standard scope-setting, clarify the primary focus on health inequality and/or health inequity, providing clear definitions and examples. This step ensures alignment with the review's objectives and establishes priorities for eligibility criteria and subsequent analysis. |
| | Frameworks and Theoretical Approach Selection | Select appropriate frameworks, such as PROGRESS Plus, incorporating both intersectional and life-course perspectives to address social determinants comprehensively. This integration enables an exploration of how health outcomes develop and vary across life stages and social contexts. |
| | Developing a Logic Model | Develop a detailed logic model that visualises potential pathways through which intersecting social and structural factors, along with life-course elements, contribute to health disparities. This model may guide data collection, analysis, and interpretation, mapping cumulative impacts on health over time. |
| Data Extraction Stage | Mapping Data | Organise and categorise data by demographic, socioeconomic, and healthcare factors to reveal disparities in access and outcomes. |
| Analysis Stage | Intersectionality | Construct an intersectionality matrix to examine how overlapping factors such as race, gender, socioeconomic status, and age shape access to care and health outcomes. Applying structured subgroup analyses can further highlight barriers faced by specific groups and trends across these intersections. |

| Stage | Component | Description |
|---|---|---|
| | Life-course Analysis | Adopt a life-course perspective by analysing health outcomes as they evolve across different life stages. Use frameworks such as the critical period model or accumulation model to assess how early-life exposures and cumulative experiences impact health trajectories over time, providing a holistic view of inequalities. |

### Supplementary Table 2: Overview of study results for medication prescribing and ethnicity

| Study ID | N participants | Outcome measure | Analysis type | Outcome data | | | | | |
|---|---|---|---|---|---|---|---|---|---|
| | | | | White/ White, non-Hispanic | Black | Hispanic/ Latino/ Latina | Asian/ Pacific Islander | Asian, other | Other |
| **Antidepressants** | | | | | | | | | |
| Allen et al., 2023 (29) | 74,309 | Use of pain medication - antidepressants | Adjusted analysis | Ref | Black/African American: OR 0.56 (95% CI 0.53 to 0.59; P < 0.05) | Hispanic: OR 0.77 (95% CI 0.73 to 0.82; P < 0.05) | Asian/Pacific Islander: 0.47 (95% CI 0.44 to 0.51; P < 0.05) | - | - |
| **Anorexia** | | | | | | | | | |
| Saphire et al., 2020 (37) | 16,246 | Any anorexia medication receipt at EOL-1 | Adjusted analysis | Ref | Black, non Hispanic: aRR 1.49 (95 CI 1.32 to 1.69; P <0.001) | Hispanic: aRR 1.13 (95% CI 0.96 to 1.33; P 0.151) | - | Asian, other: aRR 1.90 (95% CI 1.16 to 2.19; P <0.001) | - |
| **Antipsychotics** | | | | | | | | | |
| Allen et al., 2023 (29) | 74,309 | Use of pain medication - antipsychotics | Adjusted analysis | Ref | Black/African American: OR 0.54 (95% CI 0.52 to 0.57; P <0.05) | Hispanic: OR 0.74 (95% CI 0.70 to 0.78; P < 0.05) | Asian/Pacific Islander: OR 0.49 (95% CI 0.46 to 0.52; P < 0.05) | - | - |

| Study ID | N participants | Outcome measure | Analysis type | Outcome data | | | | | |
|---|---|---|---|---|---|---|---|---|---|
| | | | | White/ White, non-Hispanic | Black | Hispanic/ Latino/ Latina | Asian/ Pacific Islander | Asian, other | Other |
| | | Use of pain medication - any psychotropic medication | | Ref | Black/African American: OR 0.54 (95% CI 0.52 to 0.57; P <0.05) | Hispanic: OR 0.74 (95% CI 0.70 to 0.78; P < 0.05) | Asian/Pacific Islander: OR 0.49 (95% CI 0.46 to 0.52; P < 0.05) | - | - |
| **Anxiolytics** | | | | | | | | | |
| Allen et al., 2023 (29) | 74,309 | Use of pain medication - anxiolytics | Adjusted analysis | Ref | Black/African American: OR 0.47 (95% CI 0.43 to 0.50; P < 0.05) | Hispanic: OR 0.66 (95% CI 0.62 to 0.71; P < 0.05) | Asian/Pacific Islander: OR 0.52 (95% CI 0.48 to 0.57; P < 0.05) | - | - |
| **Appetite stimulant** | | | | | | | | | |
| Tabuyo-Martin et al., 2022 (38) | 186 | Appetite stimulant use - yes | Frequency data | White: 34 (24%) | Black: 19 (41%) P = 0.038 | - | - | - | - |
| **Dyspnoea** | | | | | | | | | |
| Saphire et al., 2020 (37) | 16,246 | Any dyspnoea medication receipt at EOL-1 | Adjusted analysis | Ref | Black, non Hispanic: aRR 0.80 (95% CI 0.70 to 0.92; P 0.002) | Hispanic: aRR 0.73 (95% CI 0.62 to 0.85; P <0.001) | - | Asian, other: aRR 0.73 (95% CI 0.63 to 0.85; P <0.001) | - |
| **Emotional distress** | | | | | | | | | |
| Saphire et al., 2020 (37) | 16,246 | Any emotional distress medication receipt at EOL-1 | Adjusted analysis | Ref | Black, non Hispanic: aRR 0.57 (95% CI 0.50 to 0.64; P <0.001) | Hispanic: aRR 0.62 (95% CI 0.53 to 0.72; P <0.001) | - | Asian, other: aRR 0.51 (95% CI 0.44 to 0.59; P <0.001) | - |

| Study ID | N participants | Outcome measure | Analysis type | Outcome data | | | | | |
|---|---|---|---|---|---|---|---|---|---|
| | | | | White/ White, non-Hispanic | Black | Hispanic/ Latino/ Latina | Asian/ Pacific Islander | Asian, other | Other |
| Tabuyo-Martin et al., 2022 (38) | 186 | Agitation medication use - yes | Frequency data | White: 77 (55%) | Black: 14 (30%) P = 0.088 | - | - | - | - |
| **Nausea/vomiting** | | | | | | | | | |
| Saphire et al., 2020 (37) | 16,246 | Any nausea/vomiting medication receipt at EOL-1 | Adjusted analysis | Ref | Black, non Hispanic: aRR 0.76 (95% CI 0.64 to 0.89; P<0.001) | Hispanic: aRR 1.01 (95% CI 0.84 to 1.22; P<0.001) | - | Asian, other: aRR 0.75 (95% CI 0.62 to 0.90; P<0.001) | - |
| Tabuyo-Martin et al., 2022 (38) | 186 | Antiemetic use - yes | Frequency data | White: 107 (76%) | Black: 35 (76%) P = 1.00 | - | - | - | - |
| **Fatigue** | | | | | | | | | |
| Saphire et al., 2020 (37) | 16,246 | Any fatigue medication receipt at EOL-1 | Adjusted analysis | Ref | Black/African American: aRR 1.48 (95% CI 1.28 to 1.72; P <0.001) | Hispanic: aRR 1.01 (95% CI 1.20 to 1.71; P <0.001) | - | Asian, other: aRR 1.38 (95% CI 1.16 to 1.63; P <0.001) | - |
| **Any pain medication (including opioids and non-opioids)** | | | | | | | | | |
| Allen et al., 2023 (29) | 74,309 | Use of pain medication – any pain medication | Adjusted analysis | Ref | Black/African American: OR 0.84 (95% CI 0.80 to 0.89; P < 0.05) | Hispanic: OR 1.07 (95% CI 1.04 to 1.17; P < 0.05) | Asian/Pacific Islander: OR 0.93 (95% CI 0.87 to 0.99; P < 0.05) | - | - |

| Study ID | N participants | Outcome measure | Analysis type | Outcome data | | | | | |
|---|---|---|---|---|---|---|---|---|---|
| | | | | White/ White, non-Hispanic | Black | Hispanic/ Latino/ Latina | Asian/ Pacific Islander | Asian, other | Other |
| | | Use of pain medication - opioids | | Ref | Black/African American: OR 0.84 (95% CI 0.79 to 0.88; P < 0.05) | Hispanic: OR 1.07 (95% CI 1.01 to 1.14; P < 0.05) | Asian/Pacific Islander: OR 0.84 (95% CI 0.79 to 0.90; P < 0.05) | - | - |
| | | Use of pain medication – non-opioids | | Ref | Black/African American: OR 0.97 (95% CI 0.90 to 1.04; P NS) | Hispanic: OR 1.16 (95% CI 1.08 to 1.25; P<0.05) | 1.37 (95% CI 1.26 to 1.49; P<0.05) | - | - |
| Enzinger et al., 2023 (31) | 318,549 | Receipt of any opioid near EOL (absolute difference in percentage points) | Adjusted analysis (model 2) | Ref | Black: -5.4% (95% CI -6.0% to -4.8%) | Hispanic: -5.4% (95% CI -6.2% to -4.6%) | - | - | - |
| | | Receipt of any long-acting opioids near EOL (absolute difference in percentage points) | | Ref | Black: -3.1% (95% CI -3.6 to -2.7%; P NR) | Hispanic: -2.4% (95% CI -3.0 to -1.9%; P NR) | - | - | - |
| | | Daily dose (morphine milligram equivalents) | | Ref | Black: -11.1 (95% CI -13.5 to -8.6; P NR) | Hispanic: -10.2 (95% CI -13.3 to -7.1; P NR) | - | - | - |

| Study ID | N participants | Outcome measure | Analysis type | Outcome data | | | | | |
|---|---|---|---|---|---|---|---|---|---|
| | | | | White/ White, non-Hispanic | Black | Hispanic/ Latino/ Latina | Asian/ Pacific Islander | Asian, other | Other |
| | | among opioid users near EOL | | | | | | | |
| | | Total dose (MMEs) filled by descendants near EOL | | Ref | Black: -237 (95% CI -269 to -207; P NR) | Hispanic: -229 (95% CI -268 to -190; P NR) | - | - | - |
| | | Urine drug screen near EOL (absolute difference in percentage points) | | Ref | Black: -0.2 (95% CI -0.5 to 0.0; P NR) | Hispanic: -1.1 (95% CI -1.5 to -0.8; P NR) | - | - | - |
| Sambamoorthi et al., 2000 (36) | 2131 | Use of pain medication in last 3 months of life | Adjusted analysis (model 3) | Ref | African-American: OR 0.63 (95% CI 0.48 to 0.83; P<0.05) | Latino/Latina: OR 0.79 (95% CI 0.56 to 1.11) | - | - | - |
| Saphire et al., 2020 (37) | 16,246 | Any pain medication at EOL-1 | Adjusted analysis | Ref | Black, non Hispanic: aRR 0.79 (95% CI 0.69 to 0.91; P 0.001) | Hispanic: aRR 0.74 (95% CI 0.63 to 0.87; P <0.001) | - | Asian, other: aRR 0.57 (95% CI 0.49 to 0.65; P <0.001) | - |
| Munir et al., 2023 (33) | 48,631 | Opioid use (1+ prescription) near EOL | Adjusted analysis | Ref | Black: OR 0.84 (95% CI 0.79 to 0.90; P <0.001) | Hispanic: OR 0.90 (95% CI 0.84 to 0.95; P <0.001) | - | Asian: OR 0.86 (95% CI 0.79 to 0.94; P <0.001) | Other: OR 0.83 (95% CI 0.74 to 0.93; P = 0.001) |

| Study ID | N participants | Outcome measure | Analysis type | Outcome data | | | | | |
|---|---|---|---|---|---|---|---|---|---|
| | | | | White/ White, non-Hispanic | Black | Hispanic/ Latino/ Latina | Asian/ Pacific Islander | Asian, other | Other |
| | | Mean daily dose near EOL (percentage difference) | | Ref | Black: -16.5% (95% CI -21.2 to -11.6%; P <0.001) | Hispanic: -19.1% (95% CI -23.5 to -14.6%; P <0.001) | - | Asian: -11.9% (95% CI -18.5 to -4.9%; P = 0.001) | Other: -7.8% (95% CI -16.4 to 1.5%; P = 0.099) |
| Rolnick et al., 2007 (35) | 421 | Likelihood of receiving high intensity pain medication 1-2 months before death | Unadjusted analysis | Ref | - | - | - | - | OR: 1.419 (95% CI 0.812 to 2.482; P = 0.219) |
| Haider et al., 2017 (32) | 750 | Morphine equivalent daily dose | Frequency data | White: median 50 (IQR 20 to 100) | Black: median 40 (IQR 25 to 80) | Hispanic: median 40 (IQR 20 to 95) | - | - | Others: median 30 (IQR 15 to 70) |
| Tabuyo-Martin et al., 2022 (38) | 186 | Analgesic use – yes | Frequency data | White: 120 (86%) | Black: 37 (80%) P = 0.482 | - | - | | |
| **Referral for palliative medicine** | | | | | | | | | |
| Tabuyo-Martin et al., 2022 (38) | 186 | Palliative medicine referral | Frequency data | n=82 White: 57 (69.5%) | n=82 Black: 25 (30.5%) P = 0.125 | - | - | - | - |
| | | No palliative medicine referral | | n=104 White: 83 (79.8%) | n=104 Black: 21 (20.2%) | | | | |

| Study ID | N participants | Outcome measure | Analysis type | Outcome data | | | | | |
|---|---|---|---|---|---|---|---|---|---|
| | | | | White/ White, non-Hispanic | Black | Hispanic/ Latino/ Latina | Asian/ Pacific Islander | Asian, other | Other |
| Abbreviations: aRR = adjusted risk ratio; CI = confidence interval; EOL = end of life; N = number; OR = odds ratio; ref = reference category | | | | | | | | | |

### Supplementary Table 3: Overview of study results for medication prescribing and gender/sex

| Study ID | N participants | Outcome measure | Analysis type | Outcome data | |
|---|---|---|---|---|---|
| | | | | Female | Male |
| **Anorexia** | | | | | |
| Saphire et al., 2020 (37) | 16,246 | Any anorexia medication receipt at EOL-1 | Adjusted analysis | aRR 0.85 (95% CI 0.79 to 0.92; P<0.001) | Ref |
| **Dyspnoea** | | | | | |
| Saphire et al., 2020 (37) | 16,246 | Any dyspnoea medication receipt at EOL-1 | Adjusted analysis | aRR 1.10 (95% CI 1.02 to 1.18; P = 0.019) | Ref |
| **Emotional distress** | | | | | |
| Saphire et al., 2020 (37) | 16,246 | Any emotional distress medication receipt at EOL-1 | Adjusted analysis | aRR 1.35 (95% CI 1.27 to 1.45; P<0.001) | Ref |
| **Fatigue** | | | | | |
| Saphire et al., 2020 (37) | 16,246 | Any fatigue medication receipt at EOL-1 | Adjusted analysis | aRR 0.89 (95% CI 0.82 to 0.97; P = 0.007) | Ref |
| **Nausea/vomiting** | | | | | |

| Study ID | N participants | Outcome measure | Analysis type | Outcome data | |
|---|---|---|---|---|---|
| | | | | Female | Male |
| Saphire et al., 2020 (37) | 16,246 | Any nausea medication receipt at EOL-1 | Adjusted analysis | aRR 1.27 (95% CI 1.17 to 1.39; P<0.001) | Ref |
| **Any pain medication** | | | | | |
| Sambamoorthi et al., 2000 (36) | 2131 | Use of pain medication in last 3 months of life | Adjusted analysis (model 3) | OR 1.44 (95% CI 1.20 to 1.74; P<0.05) | Ref |
| Enzinger et al., 2023 (31) | 318,549 | Receipt of any opioid near EOL (absolute difference in percentage points) | Adjusted analysis (model 2) | Ref | 0.7% (95% CI 0.3% to 1.0%; P <0.001) |
| | | Receipt of any long-acting opioids near EOL | | Ref | 1.1 (95% CI 0.9 to 1.4; P NR) |
| | | Daily dose (morphine milligram equivalents) among opioid users near EOL | | Ref | 9.5 (95% CI 8.1 to 11.0; P NR) |
| | | Total dose (MMEs) filled by descendants near EOL | | Ref | 140 (95% CI 122 to 158; P NR) |
| | | Urine drug screen near EOL | | Ref | 0.7 (95% CI 0.6 to 0.9; P NR) |

| Study ID | N participants | Outcome measure | Analysis type | Outcome data | |
|---|---|---|---|---|---|
| | | | | Female | Male |
| Saphire et al., 2020 (37) | 16,246 | Any pain medication at EOL-1 | Adjusted analysis | aRR 1.32 (95% CI 1.23 to 1.42; P<0.001) | Ref |
| Munir et al., 2023 (33) | 48,631 | Opioid use (1+ prescription) near EOL | Adjusted analysis | Ref | OR 0.92 (95% CI 0.89 to 0.96; P < 0.001) |
| | | Mean daily dose (MMED) near EOL (% difference) | | Ref | 2.4% (-1.1 to 6.1%; P = 0.178) |
| Haider et al., 2017 (32) | 750 | Morphine equivalent daily dose | Frequency data | Median 45 (IQR 20 to 90)<br>P = 0.042 | Median 53 (IQR 23 to 100) |
| Abbreviations: aRR = adjusted risk ratio; CI = confidence interval; EOL = end of life; N = number; OR = odds ratio; ref = reference category | | | | | |

### Supplementary Table 4: Overview of study results for medication prescribing and place of residence

| Study ID | N participants | Outcome measure | Analysis type | Outcome data | | | | | |
|---|---|---|---|---|---|---|---|---|---|
| | | | | Large metropolitan statistical area | Urban | Near urban | Rural | Less urban/ rural/ unknown Other | Elsewhere |
| **Anorexia** | | | | | | | | | |
| Saphire et al., 2020 (37) | 16,246 | Any anorexia medication receipt at EOL-1 | Adjusted analysis | Ref | aRR 0.89 (95% CI 0.76 | - | - | aRR 1.14 (95% CI 1.02 to 1.27; P = 0.018) | - |

| Study ID | N participants | Outcome measure | Analysis type | Outcome data | | | | | |
|---|---|---|---|---|---|---|---|---|---|
| | | | | Large metropolitan statistical area | Urban | Near urban | Rural | Less urban/ rural/ unknown Other | Elsewhere |
| | | | | | to 1.04; P = 0.141) | | | | |
| **Dyspnoea** | | | | | | | | | |
| Saphire et al., 2020 (37) | 16,246 | Any dyspnoea medication receipt at EOL-1 | Adjusted analysis | Ref | aRR 0.91 (95% CI 0.79 to 1.05; P = 0.196) | - | - | aRR 1.07 (95% CI 0.96 to 1.20; P = 0.212) | - |
| **Emotional distress** | | | | | | | | | |
| Saphire et al., 2020 (37) | 16,246 | Any emotional distress medication receipt at EOL-1 | Adjusted analysis | Ref | aRR 0.96 (95% CI 0.84 to 1.09; P = 0.522) | - | - | aRR 1.02 (95% CI 0.92 to 1.13; P = 0.675) | - |
| **Nausea/vomiting** | | | | | | | | | |
| Saphire et al., 2020 (37) | 16,246 | Any nausea/vomiting medication receipt at EOL-1 | Adjusted analysis | Ref | aRR 1.13 (95% CI 0.96 to 1.33; P = 0.134) | - | - | aRR 1.22 (95% CI 1.08 to 1.37; P = 0.001) | - |
| **Fatigue** | | | | | | | | | |
| Saphire et al., 2020 (37) | 16,246 | Any fatigue medication receipt at EOL-1 | Adjusted analysis | Ref | aRR 0.80 (95% CI 0.67 to 0.95; P = 0.009) | - | - | aRR 0.91 (95% CI 0.80 to 1.03; P = 0.145) | - |
| **Any pain medication** | | | | | | | | | |
| Sambamoorthi et al., 2000 (36) | 2131 | Use of pain medication in | Adjusted analysis (model 3) | - | - | OR 0.95 (95% CI 0.77 to 1.16) | - | - | Ref |

| Study ID | N participants | Outcome measure | Analysis type | Outcome data | | | | | |
|---|---|---|---|---|---|---|---|---|---|
| | | | | Large metropolitan statistical area | Urban | Near urban | Rural | Less urban/ rural/ unknown Other | Elsewhere |
| | | last 3 months of life | | | | | | | |
| Saphire et al., 2020 (37) | 16,246 | Any pain medication at EOL-1 | Adjusted analysis | Ref | aRR 1.16 (95% CI 1.01 to 1.35; P = 0.041) | - | - | aRR 1.22 (95% CI 1.09 to 1.36; P = 0.000) | - |
| Enzinger et al., 2023 (31) | 318,549 | Receipt of any opioid near EOL (absolute difference in percentage points) | Adjusted analysis | - | -3.9% (95% CI -4.3% to -3.5%; P NR) | - | Ref | - | - |
| | | Receipt of any long-acting opioids near EOL (absolute difference in percentage points) | | - | -1.9% (95% CI -2.2 to -1.6%; P NR) | - | Ref | - | - |
| | | Daily dose (morphine milligram equivalents) among opioid users near EOL | | - | -2.4% (95% CI -3.9 to -0.9%; P NR) | - | Ref | - | - |

| Study ID | N participants | Outcome measure | Analysis type | Outcome data | | | | | |
|---|---|---|---|---|---|---|---|---|---|
| | | | | Large metropolitan statistical area | Urban | Near urban | Rural | Less urban/ rural/ unknown Other | Elsewhere |
| | | Total dose (MMEs) filled by descendants near EOL | | - | -111 (95% CI -131 to -92; P NR) | - | Ref | - | - |
| | | Urine drug screen near EOL (absolute difference in percentage points) | | - | -0.4% (95% CI -0.6 to -0.2%; P NR) | - | Ref | - | - |
| Munir et al., 2023 (33) | 48,631 | Opioid use (1+ prescription) near EOL | Adjusted analysis | - | Ref | - | OR 1.13 (95% CI 1.06 to 1.20; P <0.001) | - | - |
| | | Mean daily dose (MMED) near EOL (percentage difference) | | - | Ref | - | 2.8% (95% CI -2.6 to 8.5%; P = 0.321) | - | - |

Abbreviations: aRR = adjusted risk ratio; CI = confidence interval; EOL = end of life; N = number; OR = odds ratio; ref = reference category

### Supplementary Table 5: Overview of study results for medication prescribing and personal characteristics (age)

| Study ID | N participants | Outcome measure | Analysis type | Outcome data | | | | |
|---|---|---|---|---|---|---|---|---|
| | | | | Age 66-69 | Age 70-74 | Are 75-79 | 80+ | 1 unit increase in age |
| **Anorexia** | | | | | | | | |
| Saphire et al., 2020 (37) | 16,246 | Any anorexia medication receipt at EOL-1 | Adjusted analysis | Ref | aRR 0.99 (95% CI 0.88 to 1.12; P = 0.901) | aRR 1.09 (95% CI 0.96 to 1.23; P = 0.187) | aRR 1.02 (95% CI 0.91 to 1.15; P = 0.698) | - |
| **Dyspnoea** | | | | | | | | |
| Saphire et al., 2020 (37) | 16,246 | Any dyspnoea medication receipt at EOL-1 | Adjusted analysis | Ref | aRR 1.05 (95% CI 0.93 to 1.18; P = 0.443) | aRR 0.95 (95% CI 0.85 to 1.07; P = 0.424) | aRR 0.86 (95% CI 0.77 to 0.97; P = 0.011) | - |
| **Emotional distress** | | | | | | | | |
| Saphire et al., 2020 (37) | 16,246 | Any emotional distress medication receipt at EOL-1 | Adjusted analysis | Ref | aRR 0.88 (95% CI 0.79 to 0.97; P = 0.012) | aRR 0.74 (95% CI 0.67 to 0.83; P <0.001) | aRR 0.68 (95% CI 0.61 to 0.75; P <0.001) | - |
| **Nausea/vomiting** | | | | | | | | |
| Saphire et al., 2020 (37) | 16,246 | Any nausea/vomiting medication receipt at EOL-1 | Adjusted analysis | Ref | aRR 0.88 (95% CI 0.78 to 0.99; P 0.040) | aRR 0.78 (95% CI 0.68 to 0.88; P <0.001) | aRR 0.57 (95% CI 0.50 to 0.65; P <0.001) | - |
| **Fatigue** | | | | | | | | |
| Saphire et al., 2020 (37) | 16,246 | Any fatigue medication receipt at EOL-1 | Adjusted analysis | Ref | aRR 0.92 (95% CI 0.82 to 1.04; P = 0.195) | aRR 0.78 (95% CI 0.69 to 0.89; P <0.001) | aRR 0.68 (95% CI 0.60 to 0.77; P < 0.001) | - |
| **Any pain medication** | | | | | | | | |

| Study ID | N participants | Outcome measure | Analysis type | Outcome data | | | | |
|---|---|---|---|---|---|---|---|---|
| | | | | Age 66-69 | Age 70-74 | Are 75-79 | 80+ | 1 unit increase in age |
| Saphire et al., 2020 (37) | 16,246 | Any pain medication at EOL-1 | Adjusted analysis | Ref | aRR 0.82 (95% CI 0.73 to 0.92; P = 0.001) | aRR 0.68 (95% CI0.60 to 0.76 ; P <0.001) | aRR 0.45 (95% CI 0.41 to 0.51; P < 0.001) | - |
| Munir et al., 2023 (33) | 48,631 | Opioid use (1+ prescription) near EOL | Adjusted analysis | - | - | - | - | OR 0.80 (95% CI 0.79 to 0.82; P < 0.001) |
| | | Mean daily dose (MMED) near EOL (% difference) | | - | - | - | - | -20.1% (95% CI -21.1 to -19.1%; P < 0.001) |
| Abbreviations: aRR = adjusted risk ratio; CI = confidence interval; EOL = end of life; N = number; OR = odds ratio; ref = reference category | | | | | | | | |

### Supplementary Table 6: Overview of study results for medication prescribing and personal characteristics (age at diagnosis)

| Study ID | N participants | Outcome measure | Analysis type | Outcome data | | | |
|---|---|---|---|---|---|---|---|
| | | | | Age 18-29 | Age 30-39 | Age 40-49 | Age 50 and over |
| **Any pain medication** | | | | | | | |
| Sambamoorthi et al., 2000 (36) | 2131 | Use of pain medication in last 3 months of life – waiver/race interactions | Adjusted analysis | Ref | OR 1.08 (95% CI 0.83 to 1.40; P not statistically significant) | OR 1.14 (95% CI 0.85 to 1.51; P not statistically significant) | OR 0.96 (95% CI 0.63 to 1.44; P not statistically significant) |
| Abbreviations: CI = confidence interval; N = number; OR = odds ratio; ref = reference category | | | | | | | |

### Supplementary Table 7: Overview of study results for medication prescribing and SES

| Study ID | N participants | Outcome measure | Analysis type | Outcome data |
|---|---|---|---|---|
| **Anorexia** | | | | |
| Saphire et al., 2020 (37) | 16,246 | Any anorexia medication receipt at EOL-1 | Adjusted analysis | Poverty rates (census tract levels)<br>Ref - low census tract<br>5% to <10%: aRR 0.99 (95% CI 0.88 to 1.12; P 0.931)<br>10% to <20%: aRR 1.16 (95% CI 1.04 to 1.30; P = 0.010)<br>High (20% to 100%): aRR 1.32 (95% CI 1.17 to 1.49; P<0.001) |
| **Dyspnoea** | | | | |
| Saphire et al., 2020 (37) | 16,246 | Any dyspnoea medication receipt at EOL-1 | Adjusted analysis | Poverty rates (census tract levels)<br>Ref - low census tract<br>5% to <10%: aRR 1.04 (95% CI 0.94 to 1.16; P = 0.439)<br>10% to <20%: aRR 1.03 (95% CI 0.92 to 1.14; P = 0.643)<br>High (20% to 100%): aRR 0.94 (95% CI 0.83 to 1.06; P = 0.322) |
| **Emotional distress** | | | | |
| Saphire et al., 2020 (37) | 16,246 | Any emotional distress medication receipt at EOL-1 | Adjusted analysis | Poverty rates (census tract levels)<br>Ref - low census tract<br>5% to <10%: aRR 0.94 (95% CI 0.86 to 1.04; P = 0.228)<br>10% to <20%: aRR 0.86 (95% CI 0.78 to 0.95; P = 0.003)<br>High (20% to 100%): aRR 0.80 (95% CI 0.72 to 0.90l P <0.001) |
| **Nausea/vomiting** | | | | |
| Saphire et al., 2020 (37) | 16,246 | Any nausea/vomiting medication receipt at EOL-1 | Adjusted analysis | Poverty rates (census tract levels)<br>Ref - low census tract<br>5% to <10%: aRR 0.98 (95% CI 0.87 to 1.11; P = 0.752)<br>10% to <20%: aRR 1.04 (95% CI 0.92 to 1.17; P = 0.547)<br>High (20% to 100%): aRR 0.99 (95% CI 0.87 to 1.14; P = 0.929) |
| **Fatigue** | | | | |

| Study ID | N participants | Outcome measure | Analysis type | Outcome data |
|---|---|---|---|---|
| Saphire et al., 2020 (37) | 16,246 | Any fatigue medication receipt at EOL-1 | Adjusted analysis | Poverty rates (census tract levels)<br>Ref - low census tract<br>5% to <10%: aRR 0.92 (95% CI 0.81 to 1.03; P = 0.145)<br>10% to <20%: aRR 0.86 (95% CI 0.77 to 0.97; P = 0.015)<br>High (20% to 100%): aRR 0.89 (95% CI 0.78 to 1.01; P = 0.078) |
| **Any pain medication** | | | | |
| Sambamoorthi et al., 2000 (36) | 2131 | Use of pain medication in last 3 months of life – waiver/race interactions | Adjusted analysis | African-American - ACCAP: 1.13 (95% CI 0.71 to 1.81; P not statistically significant)<br>Latino/Latina - ACCAP: 1.31 (95% CI 0.72 to 2.42; P not statistically significant) |
| | | Use of pain medication in last 3 months of life - waiver status | | Waiver status<br>Ref=Non ACCAP<br>ACCAP: 2.30 (95% CI 1.59 to 3.36; P <0.05) |
| Saphire et al., 2020 (37) | 16,246 | Any pain medication at EOL-1 | Adjusted analysis | Poverty rates (census tract levels)<br>Ref - low census tract<br>5% to <10%: aRR 1.10 (95% CI 1.00 to 1.22; P = 0.051)<br>10% to <20%: aRR 1.10 (95% CI 1.00 to 1.21; P = 0.055)<br>High (20% to 100%): aRR 1.21 (95% CI 1.08 to 1.35; P = 0.001) |
| Enzinger 2023 – low income (dually eligible for Medicare and Medicaid) | 318,549 | Receipt of any opioid near EOL (absolute difference in percentage points) | Adjusted analysis | Low income (dually eligible for Medicare and Medicaid)<br>Ref = non-dual eligible<br>Dual eligible: 6.1% (95% CI 5.7% to 6.5%; P NR) |

| Study ID | N participants | Outcome measure | Analysis type | Outcome data |
|---|---|---|---|---|
| | | Receipt of long acting opioids near EOL | | Low income (dually eligible for Medicare and Medicaid)<br>Ref = non-dual eligible<br>Dual eligible: 2.1 (95% CI 1.7 to 2.4; P NR) |
| | | Daily dose (morphine milligram equivalents) among opioid users near EOL | | Low income (dually eligible for Medicare and Medicaid)<br>Ref = non-dual eligible<br>Dual eligible: 5.8 (95% CI 4.3 to 7.4; P NR) |
| | | Total dose (morphine milligram equivalents) filled by descendants near EOL | | Low income (dually eligible for Medicare and Medicaid)<br>Ref = non-dual eligible<br>Dual eligible: 192 (95% CI 171 to 213; P NR) |
| Enzinger 2023 - community-level deprivation (SDI quintile) | 318,549 | Receipt of any opioid near EOL (absolute difference in percentage points) | Adjusted analysis | Community-level deprivation (SDI quintile)<br>Ref = Q1 lowest)<br>Q2: -0.3 (95% CI -0.8 to 0.2; P NR)<br>Q3: 0.0 (95% CI -0.5 to 0.6; P NR)<br>Q4: -0.6 (95% CI -1.1 to 0.0; P NR)<br>Q5 (highest): -1.3 (95% CI -1.9 to -0.8; P NR) |
| | | Receipt of long acting opioids near EOL | | Community-level deprivation (SDI quintile)<br>Ref = Q1 lowest<br>Q2: 0.0 (95% CI -0.4 to 0.3; P NR)<br>Q3: 0.0 (95% CI -0.5 to 0.3; P NR)<br>Q4: -0.8 (95% CI -1.1 to -0.4; P NR)<br>Q5 (highest): -1.4 (95% CI -1.8 to -1.0; P NR) |

| Study ID | N participants | Outcome measure | Analysis type | Outcome data |
|---|---|---|---|---|
| | | Daily dose (morphine milligram equivalents) among opioid users near EOL | | Community-level deprivation (SDI quintile)<br>Ref = Q1 lowest<br>Q2: 0.6 (95% CI -1.4 to 2.6; P NR)<br>Q3: -0.3 (95% CI -2.3 to 1.7; P NR)<br>Q4: -2.6 (95% CI -4.7 to -0.5; P NR)<br>Q5 (highest): -3.0 (95% CI -5.2 to -0.8; P NR) |
| | | Total dose (morphine milligram equivalents) filled by descendants near EOL | | Community-level deprivation (SDI quintile)<br>Ref = Q1 lowest<br>Q2: -0.6 (95% CI -26 to 25; P NR)<br>Q3: -4.2 (95% CI -30 to 22; P NR)<br>Q4: -41 (95% CI -68 to -14; P NR)<br>Q5 (highest): -64 (95% CI -92 to -35; P NR) |
| | | Urine drug screen near EOL | | Community-level deprivation (SDI quintile)<br>Ref = Q1 lowest<br>Q2: 0.1 (95% CI 0.1 to 0.3; P NR)<br>Q3: 0.2 (95% CI 0.0 to 0.4; P NR)<br>Q4: 0.4 (95% CI 0.1 to 0.6; P NR)<br>Q5 (highest): 0.6 (95% CI 0.4 to 0.9; P NR) |
| Munir et al., 2023 (33) – poverty duration | 48,631 | Opioid use (1+ prescription) near EOL | Adjusted analysis | Poverty duration<br>Ref=never high poverty<br>Intermittent high poverty: 1.09 (95% CI 1.02 to 1.17; P = 0.014)<br>Persistent poverty: 1.18 (95% CI 1.09 to 1.27; P < 0.001) |
| | | Mean daily dose (MMED) near EOL - % difference | | Poverty duration<br>Ref=never high poverty<br>Intermittent high poverty: -11.9% (95% CI -17.1 to -6.4%; P < 0.001)<br>Persistent poverty: -10.4% (95% CI -16.2 to -4.3%; P = 0.001) |

| Study ID | N participants | Outcome measure | Analysis type | Outcome data |
|---|---|---|---|---|
| Munir et al., 2023 (33) – dual eligibility for Medicare and Medicaid | 48,631 | Opioid use (1+ prescription) near EOL | Adjusted analysis | Dual eligibility for Medicare and Medicaid<br>Ref=non-dual<br>Dual: OR 1.15 (95% CI 1.10 to 1.21; P <0.001) |
| | | Mean daily dose (MMED) near EOL - % difference | | Dual eligibility for Medicare and Medicaid<br>Ref=non-dual<br>Dual: 5.4% (95% CI 1.3 to 7.4%; P = 0.001) |
| Abbreviations: ACCAP: HIV/AIDS specific Medicaid Home- and Community-Based Waiver Program; aRR = adjusted risk ratio; CI = confidence interval; EOL = end of life; N = number; OR = odds ratio; ref = reference category | | | | |

### Supplementary Table 8: Overview of study results for medication prescribing and social capital

| Study ID | N participants | Outcome measure | Analysis type | Outcome data | |
|---|---|---|---|---|---|
| | | | | Married | Unmarried |
| **Anorexia** | | | | | |
| Saphire et al., 2020 (37) | 16,246 | Any anorexia medication receipt at EOL-1 | Adjusted analysis | Ref | aRR 0.84 (95% CI 0.78 to 0.91; P <0.001) |
| **Dyspnoea** | | | | | |
| Saphire et al., 2020 (37) | 16,246 | Any dyspnoea medication receipt at EOL-1 | Adjusted analysis | Ref | aRR 0.92 (95% CI 0.85 to 1.00; P = 0.051) |
| **Emotional distress** | | | | | |
| Saphire et al., 2020 (37) | 16,246 | Any emotional distress | Adjusted analysis | Ref | aRR 0.92 (95% CI 0.86 to 0.99; P = 0.026) |

| Study ID | N participants | Outcome measure | Analysis type | Outcome data | |
|---|---|---|---|---|---|
| | | | | Married | Unmarried |
| | | medication receipt at EOL-1 | | | |
| **Nausea/vomiting** | | | | | |
| Saphire et al., 2020 (37) | 16,246 | Any nausea/vomiting medication receipt at EOL-1 | Adjusted analysis | Ref | aRR 0.90 (95% CI 0.83 to 0.98; P = 0.021) |
| **Fatigue** | | | | | |
| Saphire et al., 2020 (37) | 16,246 | Any fatigue medication receipt at EOL-1 | Adjusted analysis | Ref | aRR 0.96 (95% CI 0.88 to 1.05; P = 0.376) |
| **Any pain medication** | | | | | |
| Saphire et al., 2020 (37) | 16,246 | Any pain medication at EOL-1 | Adjusted analysis | Ref | aRR 0.98 (95% CI 0.91 to 1.05; P = 0.558) |
| Abbreviations: aRR = adjusted risk ratio; CI = confidence interval; EOL = end of life; N = number; ref = reference category | | | | | |
